# Artificial Intelligence Enables Quantitative Assessment of Ulcerative Colitis Histology

**DOI:** 10.1101/2022.04.28.22274339

**Authors:** Fedaa Najdawi, Kathleen Sucipto, Pratik Mistry, Stephanie Hennek, Christina Jayson, Mary Lin, Darren Fahy, Shawn Kinsey, Ilan Wapinski, Andrew H. Beck, Murray B. Resnick, Archit Khosla, Michael G. Drage

**Affiliations:** PathAI, Inc., Boston, MA, USA; PathAI Diagnostics, Memphis, TN, USA

## Abstract

Ulcerative colitis (UC) is a chronic inflammatory bowel disease that is characterized by a relapsing and remitting course. Appropriate assessment of disease activity is critical for adequate treatment decisions. In addition to endoscopic mucosal healing, histologic remission is emerging as a treatment target and a key factor in the evaluation of disease activity and therapeutic efficacy. However, there is no standardized definition of histologic remission, limiting the utility of histologic scoring, and manual pathologist evaluation is subject to intra-and inter-observer variability. Machine learning approaches are increasingly being developed to aid pathologists in accurate and reproducible scoring of histology, and can enable sensitive assessment of clinically relevant features. Here we report a proof-of-concept study using the PathAI platform to develop ML models for identification and quantification of UC histological features directly from hematoxylin and eosin (H&E)-stained whole slide images. Model-predicted histological features were used to quantify tissue area proportions and cell count proportions and densities, which correlated with disease severity and pathologist-assigned Nancy Histological Index (NHI) scores. Moreover, using multivariate analysis based on selected model-predicted histological features, we were able to accurately predict NHI scores, with a weighted kappa (k=0.93) and Spearman correlation (ρ=0.93, p<0.001) when compared to manual pathologist consensus NHI scores. We were also able to predict histological remission, based on the resolution of active inflammation, with high accuracy of 0.94. These results demonstrate the accuracy of ML models in quantifying histologic features of UC and predicting NHI scores, and highlight the potential of this approach to enable standardized and robust assessment of histologic remission for improved evaluation of disease activity and prognosis.

## Introduction

Definitions of disease remission and identification of treatment targets in inflammatory bowel disease (IBD) are multifaceted and evolving, with significant implications for clinical trial design and patient care. Assessment of disease status has evolved from the use of laboratory indices and symptoms [1] to methods that integrate clinical and endoscopic (Mayo endoscopic score (MES)) scoring to assess mucosal healing [2–4]. Recently, the histological assessment of disease activity has also been incorporated based on abundant evidence that histological active inflammation predicts increased risk of disease relapse and poor outcomes [5,6], with similar effect sizes in the context of MES 1 and 0 [5,6]. Accordingly, histologic evaluation is considered a key factor in the assessment of disease activity and therapeutic efficacy [7–10], and the cost/benefit of treating to histologic remission is currently being compared to the conventional endpoint of mucosal healing in a clinical trial (ClinicalTrials.gov ID:NCT04259138). In its draft guidance to industry on UC clinical trial endpoints, the US Food and Drug Administration proposed that label claims of mucosal healing be supported by both endoscopic and histologic assessments [11,12]. However, the FDA has not established acceptance of a standardized system for histologic assessments.

An international panel of IBD gastroenterologists and pathologists has recommended that the Geboes Score (GS), the Robarts Histopathology Index (RHI), and/or the Nancy Histological Index (NHI) be used to assess histologic activity in UC clinical trials, and the European Crohn’s and Colitis organization recommends the RHI and NHI [10,13]. A retrospective analysis of biopsies collected in a phase 2 clinical trial for ozanimod treatment found that RHI, GS, and NHI scores by expert GI pathologists, were similarly reliable and able to assess histological changes resulting from treatment [14]. However, manual histologic scoring is subject to variability in intra-pathologist and inter-pathologist evaluation, and is time consuming. In a study of pathologist evaluation of histological remission in UC biopsies, concordance between general pathologist and GI-specialist pathologist scores was poor [15,16]. Suboptimal inter-pathologist concordance was also observed in the evaluation of cohorts enriched for mild disease activity [17]; a pattern also observed in endoscopic assessment of mucosal healing [18]. A robust standardized tool to assess histopathological disease activity that is reproducible across the full spectrum of disease would standardize objective measures of disease activity and facilitate inter-trial comparisons.

Machine learning (ML)-based evaluation of digitized whole slide images (WSI) of human tissue has shown remarkable promise to enable precise quantitative assessment of histopathology, functioning both as an assistive device to reduce intra- and inter-observer variability, and provide rich data amenable to powerful statistical analyses [19]. ML-based models can be trained to segment regions of tissue and identify cell types across WSI and support pathologist scoring by providing accurate, quantitative, and scalable assessment of tissue samples [20].

Here we report ML-based models to quantify UC histological features in WSI of H&E-stained colon biopsies, which are used as input to predict consensus slide-level NHI scores. This ML-based assessment of UC histological features and disease severity is a step towards robust, reproducible, and quantitative characterization of UC histology to better guide patient treatment decisions and standardize histological endpoint definitions.

## Methods

### Data set characteristics

Models were developed using 637 digitized whole slide images (WSI) of hematoxylin and eosin (H&E)-stained left colon and rectum biopsies from 334 adult patients with various histologic patterns: normal colon (n=123), active colitis (n=17), inactive chronic colitis (n=154), or chronic colitis with mild (n=142), moderate (n=117), or severe (n=84) histologic activity. The slides were scanned at 40x objective magnification using Aperio GT450 (383 slides) and Aperio AT2 (254 slides) scanners. The slides were then split into training (490; 77%) and validation (147; 23%). An additional 293 held-out slides from the PathAI Diagnostics laboratory were used for evaluation of model performance. These slides were scanned with 40x objective magnification using Aperio GT450 (212 slides) and Aperio AT2 (81 slides), and were comprised of slides representing normal colon (n=17), inactive chronic colitis (n=123), and chronic colitis with mild (n=51), moderate (n=50), or severe (n=52) histologic activity.

### Machine learning-based model development

WSI were annotated by American Board of Pathology board-certified pathologists with expertise in gastrointestinal pathology. In total, ∼38,000 tissue annotations and ∼124,000 cell annotations were collected. Using these annotations, the PathAI research platform was then applied to train a convolutional neural network with over 20 layers and 8 million parameters to produce pixel-level predictions of ulcerative colitis histology.

Tissue region models were trained to segment areas of normal epithelium, neutrophil infiltrated epithelium, goblet cell cytoplasm, crypt abscess, inter-gland lumen, blood vessels, lamina propria, muscularis mucosa, basal plasmacytosis, erosion/ulceration, and granulation tissue. The models were also trained to recognize submucosal tissue and lymphoid aggregates (regardless of location), which were excluded from analysis. Cell labeling models were trained to identify neutrophils, plasma cells, intraepithelial lymphocytes, non-intraepithelial lymphocytes, eosinophils, goblet cell nuclei, and enterocytes. A separate class called “other cells” was provided to include cells other than the ones previously mentioned; this includes but is not limited to endothelial cells and smooth muscle cells. Tissue and cell model predictions were then visualized as colored overlays on the WSI.

To enhance model generalizability, we performed image augmentation by introducing varied brightness and contrast and altered image orientations. Model development was carried out in iterative cycles involving annotation collection, annotation quality control, model training, and performance review, and the best performing models were selected by tracking qualitative and quantitative performance metrics (accuracy, F1 score, loss values) across model iterations.

### Slide background and artifact exclusion

Most pixels in a whole slide image of colon biopsies contain background rather than tissue, and tissue-containing regions may contain sample preparation and imaging artifacts, including debris, tissue folds, and areas of poor focus. We excluded both background and artifact-containing regions from our analysis using an additional convolutional neural network (CNN) trained on multiple organs, which involved the existing tissue annotations and an additional ∼10,000 artifact and background annotations to classify pixels as either background, tissue with artifact, or usable tissue. Slides with a high proportion of artifact and background were manually reviewed by pathologists, and if deemed uninterpretable, were removed from the training dataset. All other models and features were then evaluated only in the areas classified as usable tissue.

### Evaluation of model predictions of UC histological features

PathAI internal pathologists (FN, MD and MR) performed qualitative review of tissue and cell overlays representing model predictions on H&E images for any misclassifications by the model. This qualitative review helped guide the iterative model development.

To establish ground-truth for the cell model prediction accuracy, we selected image patches ‘frames’ (320 × 320 pixels, ∼80 × 80 *μ*m for 40x scanned slides; n=160) across different cell densities and tissue regions to ensure a representation of a range of cell distributions for each cell type. Exhaustive annotations were then collected from five pathologists to produce quantitative estimates of cell identity for epithelial cells, lymphocytes, plasma cells, eosinophils and neutrophils. After the annotations were collected, hierarchical clustering was performed on the pathologists’ annotations and the model cell detection results in each frame to identify cell locations; the cells detected by our model were included to resolve the pathologists’ undercalling issue. The cluster radius threshold for each frame is defined by the average of five minimum annotation distances by each pathologist to account for different cell types and sizes in each frame. Clusters with only one annotation or predicted cell are excluded. To account for the variability and bias in pathologist annotations, we used a Bayesian approach [21] to estimate the true type of each cell using the pathologists annotations as the input (**Supplementary Figure 1**). We then used these estimated ground truths to quantify the overall performance of the annotators and our model.

### Pathologist scoring of digitized WSI

Digital WSI (n=418) were scored manually using the NHI system according to published guidance [22] by five GI-fellowship trained surgical pathologists. Of these three were internal GI-fellowship trained pathologists (FN, MD, MR), and two additional GI-fellowship trained pathologists were selected for manual scoring after exhibiting proficiency in use of the Nancy Histological Index, defined as a weighted kappa score > 0.80 concordance with the consensus of the internal GI pathologists’ scores. Consensus was defined as agreement between at least three out of five of pathologists. Slides that did not achieve consensus (n=8 (1%)) were excluded, leaving 410 WSI. Out of these WSI, 117 slides were excluded in our model-derived features evaluation as they were included in the training and validation set used in the model development.

### Evaluation of model-derived features based on pathologist NHI scores

Model predictions were used to generate >800 quantitative human-interpretable features (HIFs), measuring tissue areas, area proportions (e.g., area proportion of erosion/ulceration in all usable tissue), cell counts, cell count proportions (e.g., count proportion of neutrophils in all cells in lamina propria), and cell densities (e.g., density of neutrophils in lamina propria). The correlation of each model-generated human interpretable feature with pathologist-assessed slide level NHI scores (n=293) was reported using Spearman correlation. Additionally, the Mann-Whitney U test was performed on the same set of slides to assess the distribution differences between two adjacent NHI scores for each feature.

### Prediction of NHI scores by multivariate classification models

To further assess the capability of our model, we trained multivariate classification models on the slides with assigned NHI scores (n=410), using model-derived HIFs to predict NHI. The 410 slides were further split into training (237; 60%), validation (93; 23%), and held-out test (80; 17%) set. We used 17 UC histological HIFs that were hand-picked by pathologists in the training. The multivariate classification models included logistic regression, gradient boosting, and random forest, and were run using the scikit-learn library [23]. The most optimal model based on the produced weighted kappa scores was selected using 5-fold cross validation. Finally, the performance on the most optimal model was further evaluated on the test set.

## Results

### ML model development for quantitation of UC histological features

To develop a quantitative and reproducible measure of UC histology, we trained deep CNN models to predict pathological features of UC disease severity using pathologist annotations on digitized WSI of H&E-stained samples from individuals with normal colon, chronic inactive colitis, and mild, moderate, or severe chronic active ulcerative colitis (**Figure 1**). Model predictions were visualized as colored overlays on the WSI, allowing for qualitative review of model performance. Tissue model overlays show predicted areas of normal epithelium, neutrophil infiltrated epithelium, goblet cell cytoplasm, crypt abscess, inter-gland lumen, blood vessels, lamina propria, muscularis mucosa, basal plasmacytosis, erosion/ulceration, and granulation tissue (**Figure 2**). Cell model overlays show predictions of neutrophils, plasma cells, intraepithelial lymphocytes, non-intraepithelial lymphocytes, eosinophils, goblet cell nuclei, and enterocytes (**Figure 3**).

**Figure 1.**
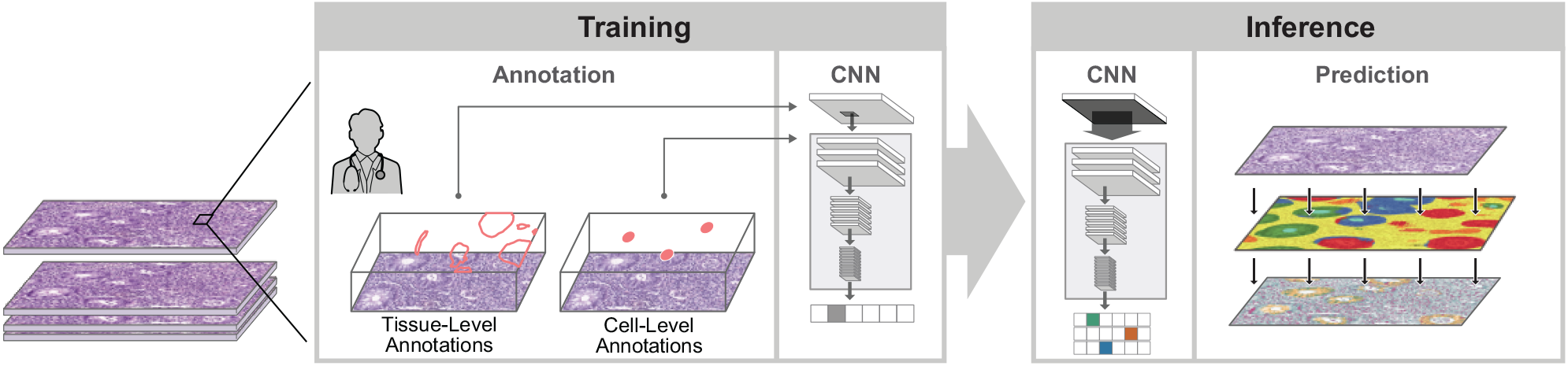
Schematic of model development for identification of UC histological features. Convolutional neural network (CNN)-based ML models were trained using manual annotations provided by pathologists identifying histological features relevant to UC. Inference of study samples generates tissue- and cell-level predictions.

**Figure 2.**
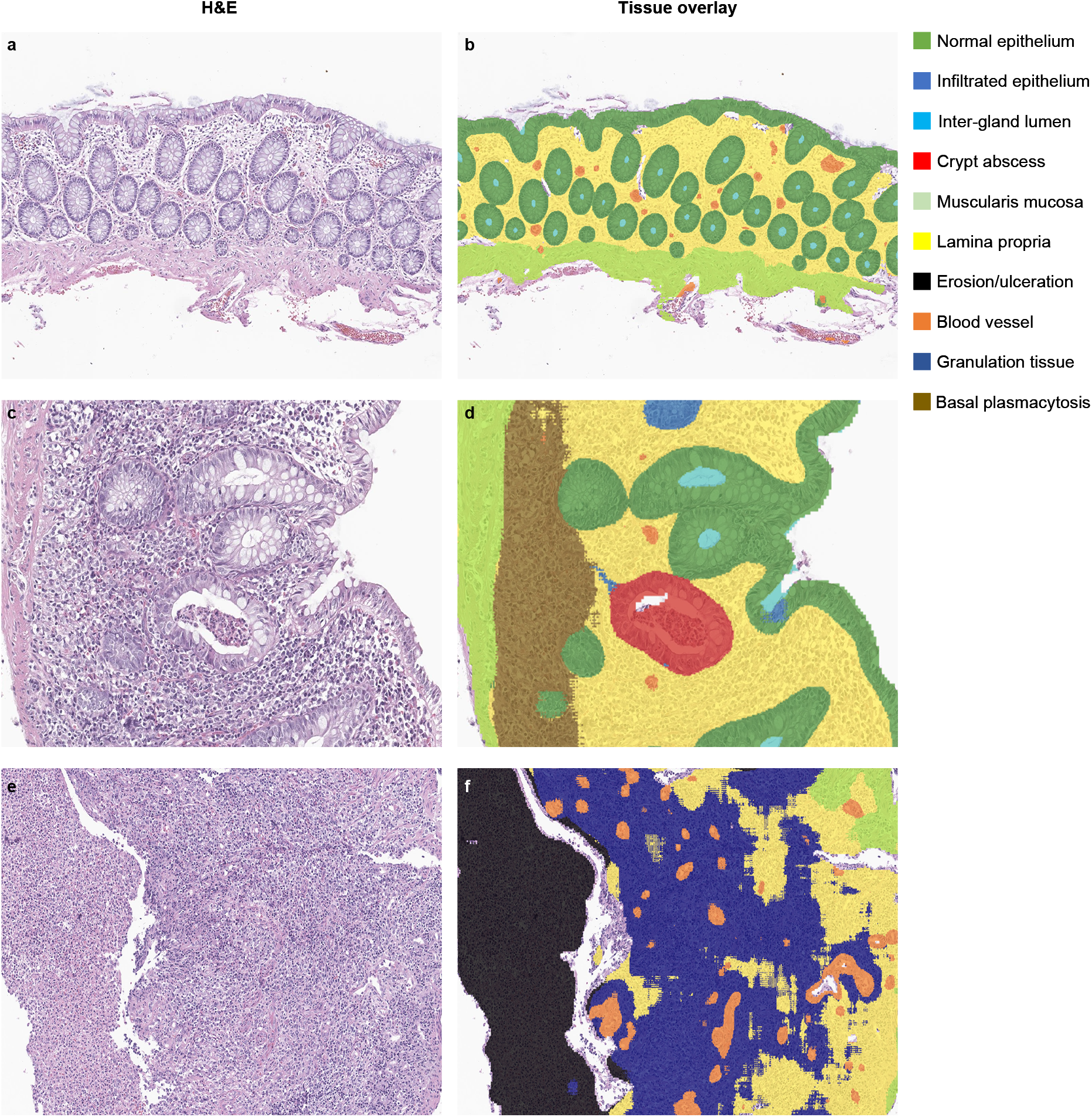
Representative fields from colon biopsies showing H&E image and corresponding model-generated tissue overlay. (**a**,**b**) Images from normal colon (NHI 0, 4x magnification). (**c**,**d**) Images from moderate chronic active colitis (NHI 3, 10x magnification). (**e**,**f**) Images from severe chronic active colitis (NHI 4, 10x magnification).

**Figure 3.**
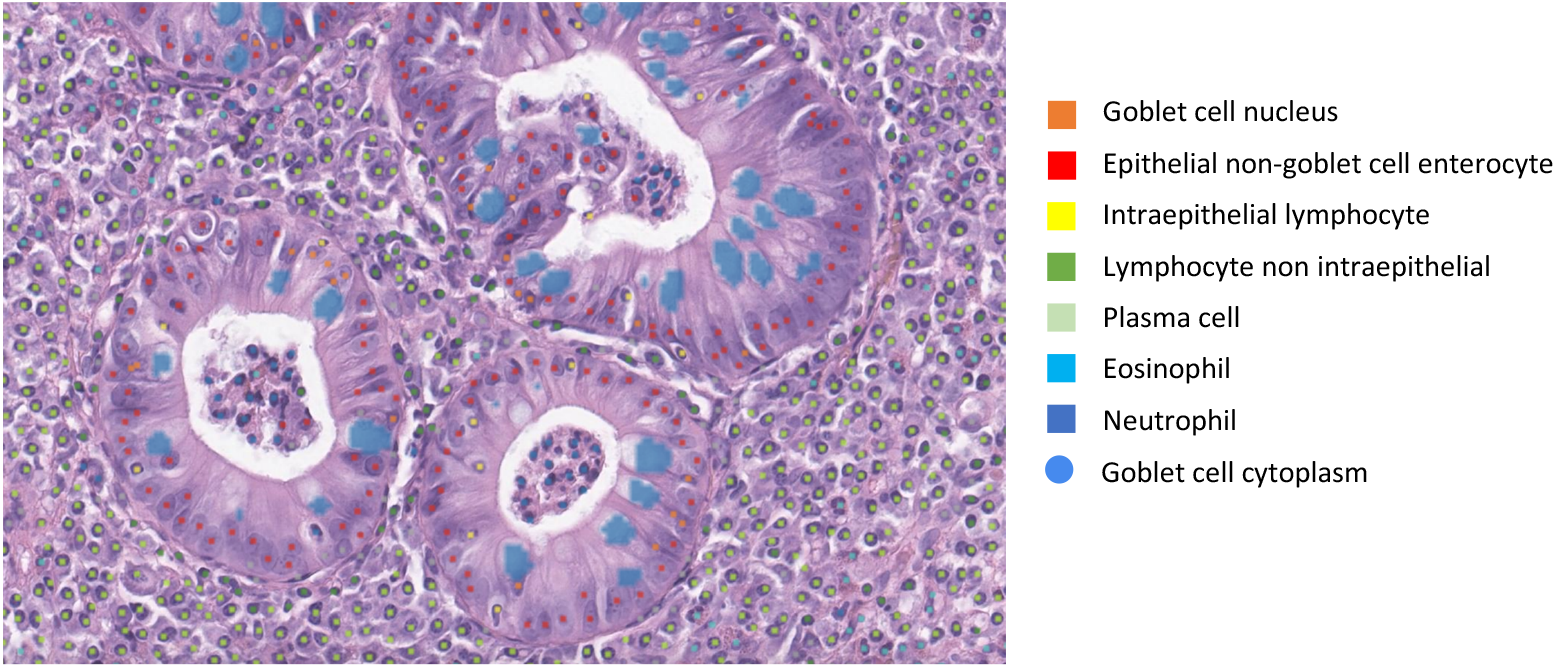
Representative field showing model-generated cell overlay. Image from moderate chronic active colitis (NHI 3, 40x magnification).

To validate our cell model predictions, we compared the performance of our model to that of the pathologists using the Bayesian-derived estimated ground truths on the 160 ∼80 × 80 *μ*m frames. Weighted average of precision and recall (F1 score), which takes both false positives and false negatives into account, was calculated for the cell model predictions and each pathologist annotation compared to consensus. The model performance was either at level with the mean or was within 1 standard deviation of the mean seen for individual pathologists across the 5 cell types: neutrophils, eosinophils, lymphocytes (intraepithelial and non-intraepithelial lymphocytes), plasma cells, and epithelial cells (goblet cells and enterocytes) (**Figure 4** and **Supplementary Figure 2**).

**Figure 4.**
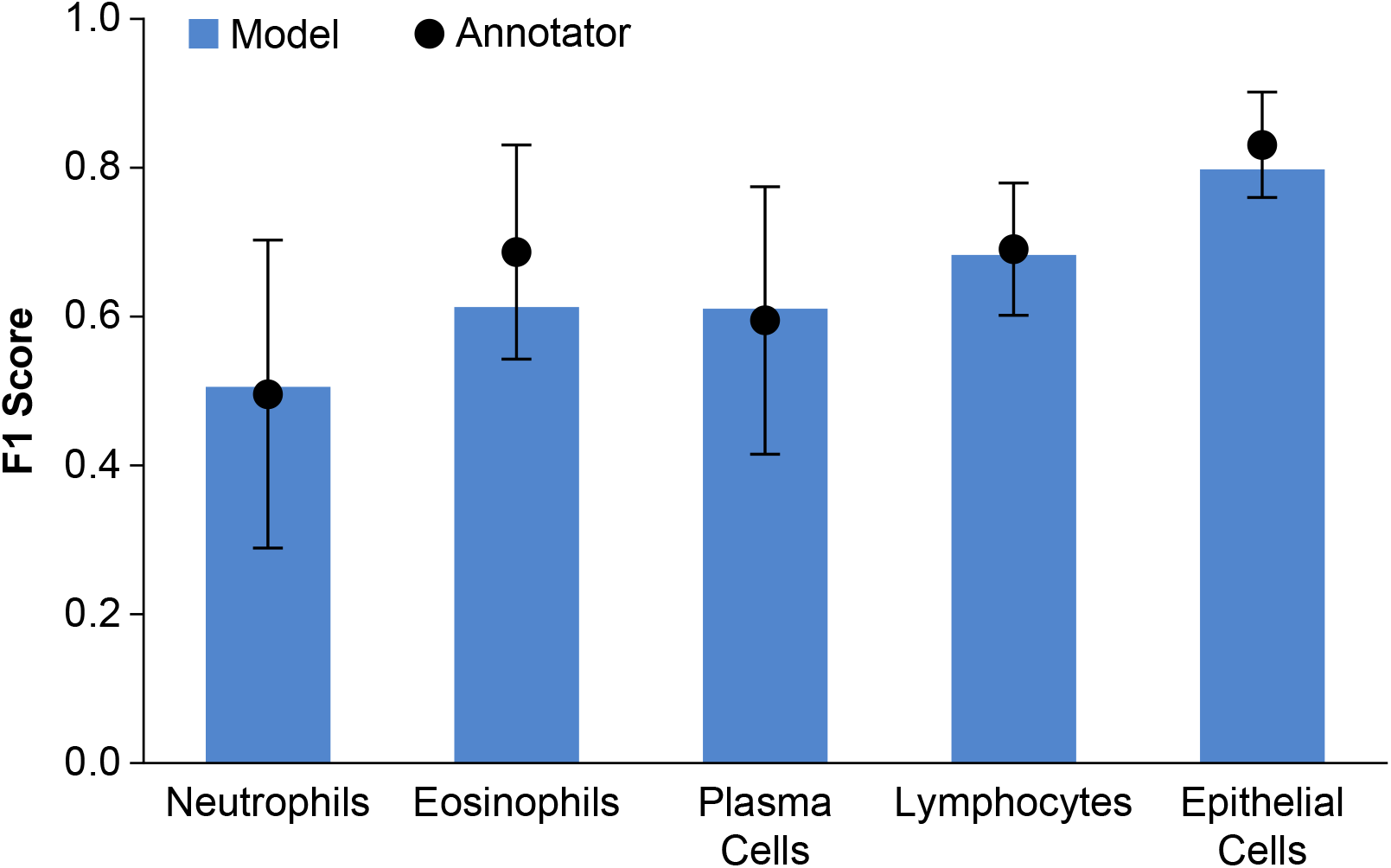
Validation of model performance. Model predictions were compared to exhaustive annotations from five pathologists on the image frames (n=160). Variability and bias in pathologist annotations were accounted for using a Bayesian approach. The weighted average of precision and recall is reported for both the cell model predictions (F1 score) and pathologists’ annotations (average F1 score) compared to consensus for each of 5 cell classes. The error bars represent the standard deviation across annotators.

### Correlation of ML-model quantitation of UC histological features with manual pathologist scoring

#### Correlation with overall disease severity (NHI 0-4)

We assessed correlation between model-generated quantitative features and pathologist consensus NHI scores. We found that quantitative HIFs are indicative of disease severity and correlated with pathologist consensus NHI scores (**Table 1**). Features that showed the strongest association are those reflective of active inflammation and epithelial injury, such as combined area proportion of infiltrated epithelium (neutrophilic infiltration), crypt abscess, erosion, ulceration and associated granulation tissue over mucosa (ρ=0.89, p<0.001), area proportion of epithelium with neutrophilic infiltration over all epithelium (ρ=0.88, p<0.001), and both count proportion and density of neutrophils in epithelium, mucosa, ulcer or their combination (ρ=range 0.78-0.89, all comparisons p<0.001) (**Figure 5**). These features show strong correlation with increasing NHI score, highlighting the relevance of the quantitative predictions made by the ML model. Count proportion of eosinophils in mucosa did not show correlation with neutrophilic inflammation (NHI 2-4) (ρ=0.1, p= 0.38), consistent with some, but not all, of the previous literature [24,25].

**Figure 5.**
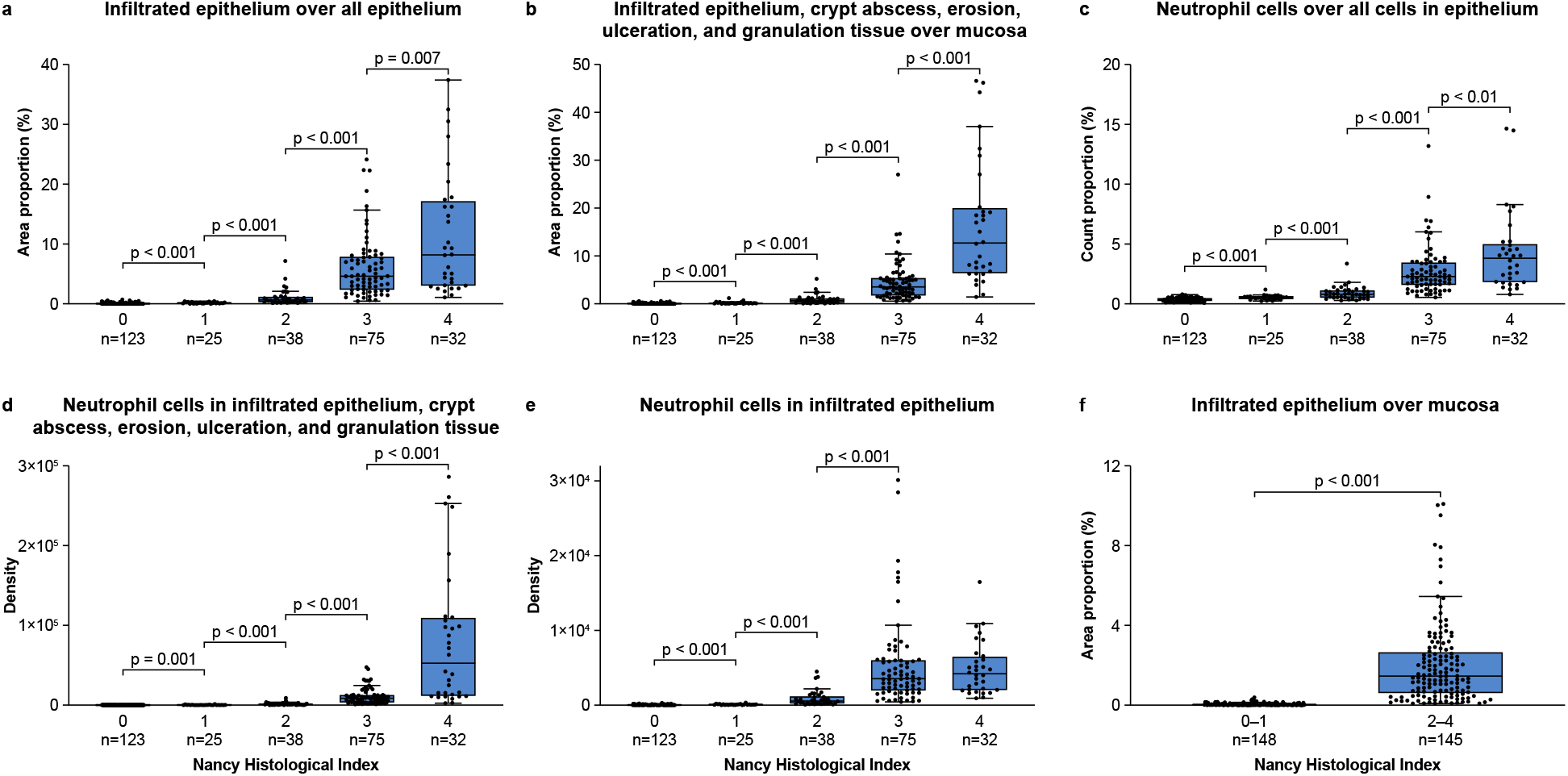
Example ML-generated features showing correlation with individual and grouped NHI scores. Quantitative HIFs measuring tissue area proportions, cell counts, and cell densities show correlation with consensus manual pathologist NHI scores.

**Table 1.** ML-generated feature correlation with pathologist NHI scores. Darker colors represent stronger correlations or lower p-values.

| FEATURES | CORRELATION | P-VALUE |
| --- | --- | --- |
| AREA PROPORTION OF INFILTRATED EPITHELIUM, CRYPT ABSCESS, EROSION OR ULCERATION, AND GRANULATION TISSUE OVER MUCOSA | 0.889 | <0.001 |
| AREA PROPORTION OF INFILTRATED EPITHELIUM, CRYPT ABSCESS, EROSION OR ULCERATION, GRANULATION TISSUE, AND BASAL PLASMACYTOSIS OVER MUCOSA | 0.885 | <0.001 |
| AREA PROPORTION OF ALL INFILTRATED EPITHELIUM AND CRYPT ABSCESS OVER ALL EPITHELIUM AND CRYPT ABSCESS | 0.883 | <0.001 |
| AREA PROPORTION OF ALL INFILTRATED EPITHELIUM OVER ALL EPITHELIUM | 0.881 | <0.001 |
| AREA PROPORTION OF ALL INFILTRATED EPITHELIUM OVER MUCOSA | 0.871 | <0.001 |
| AREA PROPORTION OF ALL INFILTRATED EPITHELIUM AND CRYPT ABSCESS OVER MUCOSA | 0.871 | <0.001 |
| AREA PROPORTION OF CRYPT ABSCESS, EROSION OR ULCERATION, AND GRANULATION TISSUE OVER MUCOSA | 0.826 | <0.001 |
| AREA PROPORTION OF GRANULATION TISSUE OVER MUCOSA | 0.822 | <0.001 |
| AREA PROPORTION OF EROSION OR ULCERATION AND GRANULATION TISSUE OVER MUCOSA | 0.79 | <0.001 |
| AREA PROPORTION OF CRYPT ABSCESS OVER ALL EPITHELIUM AND CRYPT ABSCESS | 0.777 | <0.001 |
| AREA PROPORTION OF CRYPT ABSCESS OVER MUCOSA | 0.766 | <0.001 |
| AREA PROPORTION OF EROSION OR ULCERATION OVER MUCOSA | 0.743 | <0.001 |
| AREA PROPORTION OF BASAL PLASMACYTOSIS OVER MUCOSA | 0.721 | <0.001 |
| AREA PROPORTION OF GOBLET CELL CYTOPLASM OVER ALL EPITHELIUM | -0.526 | <0.001 |
| AREA PROPORTION OF BLOOD VESSEL OVER MUCOSA | 0.42 | <0.001 |
| COUNT PROPORTION OF NEUTROPHILS OVER ALL CELLS IN ALL EPITHELIUM | 0.856 | <0.001 |
| COUNT PROPORTION OF NEUTROPHILS OVER ALL CELLS IN MUCOSA | 0.734 | <0.001 |
| COUNT PROPORTION OF LYMPHOCYTES, PLASMA CELLS, AND EOSINOPHILS OVER ALL CELLS IN MUCOSA | 0.692 | <0.001 |
| COUNT PROPORTION OF PLASMA CELLS OVER ALL CELLS IN BASAL PLASMACYTOSIS | 0.676 | <0.001 |
| COUNT PROPORTION OF LYMPHOCYTES AND PLASMA CELLS OVER ALL CELLS IN MUCOSA | 0.668 | <0.001 |
| COUNT PROPORTION OF PLASMA CELLS OVER ALL CELLS IN MUCOSA | 0.555 | <0.001 |
| COUNT PROPORTION OF NEUTROPHILS OVER ALL CELLS IN LAMINA PROPRIA | 0.511 | <0.001 |
| COUNT PROPORTION OF EOSINOPHIL CELLS OVER ALL CELLS IN MUCOSA | 0.407 | <0.001 |
| COUNT PROPORTION OF INTRAEPITHELIAL LYMPHOCYTES OVER ALL CELLS IN EPITHELIUM | 0.325 | <0.001 |
| COUNT PROPORTION OF NON-INTRAEPITHELIAL LYMPHOCYTES OVER ALL CELLS IN LAMINA PROPRIA | 0.106 | 0.071 |
| COUNT PROPORTION OF EOSINOPHIL CELLS OVER ALL CELLS IN LAMINA PROPRIA | 0.097 | 0.097 |
| DENSITY OF NEUTROPHILS IN INFILTRATED EPITHELIUM, CRYPT ABSCESS, EROSION OR ULCERATION, AND GRANULATION TISSUE | 0.892 | <0.001 |
| DENSITY OF NEUTROPHILS IN INFILTRATED EPITHELIUM | 0.879 | <0.001 |
| DENSITY OF NEUTROPHILS IN LAMINA PROPRIA | 0.712 | <0.001 |
| DENSITY OF EOSINOPHILS IN MUCOSA | 0.549 | <0.001 |

#### Correlation with severe active colitis (NHI 3,4)

Quantitative analysis of features associated with severe disease including erosion, ulceration and granulation tissue in the mucosa, showed a strong positive correlation across all NHI scores (<=0.79 p<0.001) and, as expected, significantly discriminated cases with NHI score 3 vs score 4 (p<0.001). Neutrophil density within infiltrated epithelium, crypt abscess, erosion, ulceration and granulation tissue, also produced a strong positive correlation across all NHI scores (<=0.89 p<0.001) and significantly discriminated cases with NHI score 3 from NHI score 4 (p<0.001) (**Fig. 5d**).

#### Correlation with chronic inactive colitis (NHI 0,1)

When evaluating quantitative features that distinguished lower NHI scores (NHI 0 from NHI1), as expected, we noted features related to chronic inflammation, including basal plasmacytosis area proportion, and combined count proportions of chronic inflammatory cells (plasma cells, lymphocytes and eosinophils) (both comparisons p<0.001). Interestingly, when examining quantitative features of individual cell types, we found that plasma cell and eosinophil count proportions and cell densities maintained that correlation (with plasma cells showing stronger correlation), while lymphocyte cell features within lamina propria did not differentiate NHI 0 from NHI 1 (p=0.1).

#### Correlation with grouped NHI scores

We also examined correlation of features with grouped NHI scores: NHI 0-1 vs. NHI 2-4 to reflect the cutoff for resolution of active inflammation, and NHI 0-1 vs NHI 2 vs NHI 2-4 to compare features between cases with inactive vs mild vs. moderate to severe activity. As expected, cell features related to neutrophils and resulting tissue features of active inflammation showed the strongest correlation (**Fig. 5f**).

#### Additional features

Notably, some features that showed correlation with the overall NHI score are either not components of the NHI scoring index, or are only used in a limited fashion for assigning the score of NHI 1. Such features include plasma cell features, such as basal plasmacytosis area proportion as well as plasma cell count and density within mucosa and basal plasmacytosis, which correlated with the increasing overall NHI score (**Table 1**). Other such features include goblet cell cytoplasm mucin area proportion in the epithelium, which revealed a moderate negative correlation with increasing NHI score (ρ=-0.52, p<0.001). This feature discriminated biopsies with no activity (NHI 0-1) from those with mild active disease (NHI 2) and biopsies showing more advanced disease (NHI 3-4) (ρ=-0.53, p<0.001).

### ML prediction of Nancy Histological Index (NHI)

To examine the reliability and reproducibility of manual pathology-based scoring of UC activity, five GI fellowship–trained pathologists independently graded UC activity using the NHI scoring index. We found that pathologist scoring showed near perfect interobserver reproducibility for overall score (weighted kappa average 0.94).

We then compared manual pathologist NHI scoring with ML-based NHI scoring. Multivariate analysis was performed to predict the NHI score using 17 selected histologic features (**Supplementary Figure 3**). Among all the multivariate models, a random forest classifier was selected based on the performance of a 5-fold cross validation measured by weighted kappa.

The random forest classifier model yielded a weighted kappa (k= 0.93) and Spearman correlation (ρ=0.93, p<0.001) when compared to manual pathologist consensus NHI score **(Fig. 6a**).

**Figure 6.**
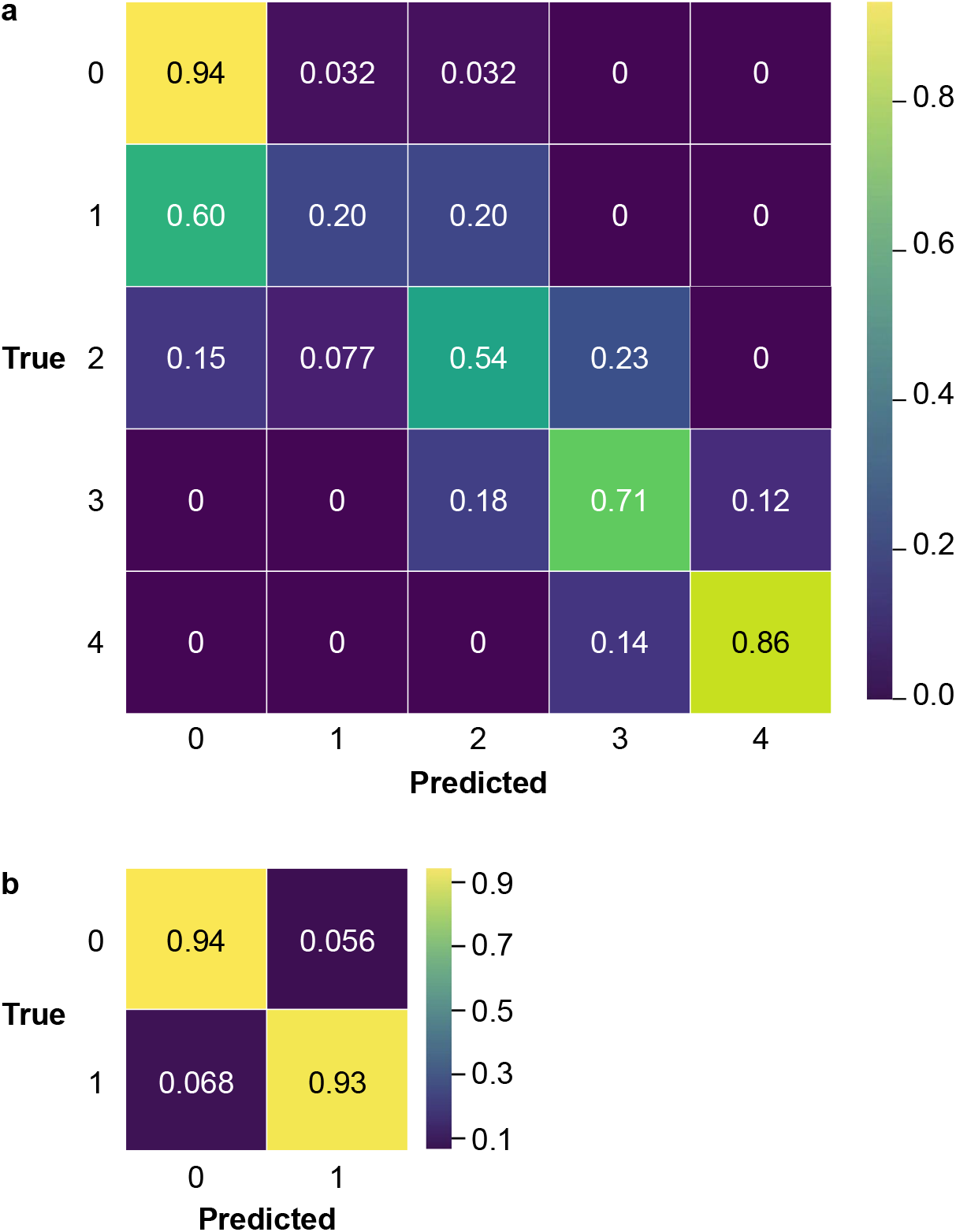
Prediction of NHI using random forest classifier trained with model-generated histologic features as input. **(a**,**b)** Confusion matrices for random forest classifier predictions compared to manual pathologist consensus NHI scores are shown for prediction of overall NHI (**a**) and for binarized prediction of chronic active colitis NHI 2-4 vs chronic inactive colitis and normal NHI 0-1 (**b**).

Notably, confusion of the model was greatest for scores with lower concordance among pathologists. The model performance differentiating NHI 0-1 and NHI 1-2 (kappa 0.32 and 0.36 respectively), although low, was within the range calculated for the 5 expert gastrointestinal pathologists (kappa ranges 0.257–0.893 and 0.308–0.806, with averages of 0.56 and 0.51, respectively).

In order to assess the model’s ability to classify histologic remission [15], we also trained a random forest classifier model to predict cases with disease activity (chronic active colitis NHI 2-4) from cases with chronic inactive colitis or those that have normal or mildly increased lamina propria density (NHI 0-1). The random forest classifier model yielded an accuracy of 0.94 (weighted kappa k= 0.87 and Spearman correlation (ρ=0.79 p<0.001) **Fig. 6b**). The model performance was within the range of agreement seen between an individual expert pathologist and consensus (kappa range 0.795–0.949), indicating that the model performed within the range of expert review, while at the same time enabling a level of reproducibility superior to what could be achieved by manual pathologist scoring.

## Discussion

Histopathology is an essential component of disease activity assessment for patients with UC. We now see a shift to routinely incorporate biopsy collection and histologic read out for most UC drug development programs. Here we report an ML-based approach to automate and standardize histological assessment of UC, and quantify key HIFs that robustly predict slide-level scores of expert pathologists according to the NHI. A random-forest classifier predicted slide-level NHI scores on a held-out test set with a high degree of accuracy and utilized HIFs in a manner similar to pathologists, further confirming the validity of this approach.

Our HIF-based approach has several unique strengths pertinent to challenges in incorporating UC histological evaluation into clinical practice and care of patients with IBD. Model predictions are visualized as overlays directly on whole slide images, providing exceptional explainability with real-time qualitative evaluation of performance. We demonstrated that model-quantified histological disease activity measures showed high correlation with consensus NHI scores of expert gastrointestinal pathologists. ML model-generated quantitative features of disease activity that reflect the natural progression of active inflammation and epithelial injury, including epithelium with neutrophilic infiltration, crypt abscess, erosion, ulceration and associated granulation tissue, as well as neutrophil cell composition within these different tissue compartments, showed strong correlation with increased disease severity, both when examined individually and in combination. These features also discriminated between cases with no active inflammation and cases with mild activity and cases showing moderate to severe disease, highlighting the robustness of our model performance.

We were able to use model-derived histologic features to directly predict NHI scores (weighted k=0.93, Spearman correlation r=0.93). This approach was also able to predict active disease, distinguishing chronic active colitis (NHI 2-4) vs. normal and chronic inactive colitis (NHI 0-1) (weighted k=0.87, Spearman correlation r=0.87).

Extraction of quantitative features from our ML-based models also enabled evaluation of histopathologic features not included in the NHI. For example, our quantitative analysis showed that both area proportion and plasma cell composition of basal plasmacytosis are correlated with increasing disease severity reflected by increased NHI score. Features quantifying plasma cells in the mucosa also correlated with increasing disease severity, highlighting the role of plasma cells in UC disease biology and progression [26]. We were also able to quantify goblet cell mucin depletion, a known feature of epithelial injury in UC that has not previously been quantified [27]. We show that goblet cell mucin is inversely correlated with increased disease severity and higher NHI scores. Other HIFs may be leveraged to prognosticate [28] and/or predict efficacy of specific treatments [24] or gain novel insights into disease biology.

Limitations of this study include the fact that the slides used to evaluate the model were from a single histology laboratory, which incurs a risk of overfitting, and does not address the question of generalizability. This risk was mitigated by the inclusion of slides scanned from two different scanners in all model training, data augmentation incorporated in the CNN model training, 5-fold cross-validation in the multivariate model selection, and usage of held-out test set for the final assessment of the RFC model. The paucity of patient metadata limits the scope of this work to prediction of histological score. Amongst histological scores, the model showed lowest performance in discriminating NHI 0-1, likely due to a combination of limited data in NHI 1, heterogeneity across the tissue fragments on the slide, and the higher interobserver variability among pathologists in assigning a score of NHI 1. We compared our RFC predictions to the majority consensus scores of a selected group of subspeciality trained pathologists with near perfect ICC. Since histological scoring is likely to be incorporated into routine diagnostic practice [29], future ‘real world’ studies are needed to determine relative performance of the model amongst a greater number of pathologists of broader background and experience. Ongoing work will allow prediction of additional histological scoring systems, and explore associations with clinical metadata.

We envision AI-assisted histological analysis as an integral component of how big data will contribute to precision medicine for patients with IBD [30], analogous to its established role in advancing oncology research, often demonstrating the ability to predict clinically relevant molecular features, refine prognostic information, guide therapeutic selection, or predict gene expression signatures from H&E images [31], and allow rich data extraction from H&E images [32]. In the immediate future, explainable AI can function as an assist device with the potential to reduce inter-observer variability, facilitate decentralized trials, or could serve as an automated predictor equivalent to a centralized reader, similar to the demonstrated utility in evaluation of endoscopic images [33–35]. This has particular relevance in mild disease, where inter-observer variability is greatest. The HIF outputs can easily be combined with orthogonal data streams (clinical metadata, gene expression profiling of mucosal biopsies, spatial transcriptomics), and the combination is expected to demonstrate powerful synergy.

We report a proof-of-concept study demonstrating that machine learning models can quantify histologic features of UC from H&E images, and that quantitative model-generated HIFs predict NHI scores with high accuracy at the threshold of ‘histologic remission’ (NHI 0-1) suggested by a consensus of experts [15]. While the scope of this current study is limited to predicting NHI scores, we believe that this ML-based assessment of UC histological disease severity is a stepping stone toward a robust and reproducible ML-based image analysis for quantitative characterization of UC histology, with tremendous potential as a tool for measuring disease severity, stratifying risk, monitoring treatment response, and ultimately advancing the care of patients with inflammatory bowel disease.

## Data Availability

All data produced in the present work are contained in the manuscript.

