## Supplementary Figures for "Artificial Intelligence Enables Quantitative Assessment of Ulcerative Colitis Histology"

### Supplementary Information

**Supplementary Figure 1. The Bayesian-based cell frame analysis pipeline.** Hierarchical clustering was first performed on all pathologists' annotations and the model cell detection results to locate true cells. For each detected true cell, we ran a Bayesian model with the pathologists' annotations only as the input to estimate the cell type ground truth based on the estimated annotators' specificity and sensitivity.

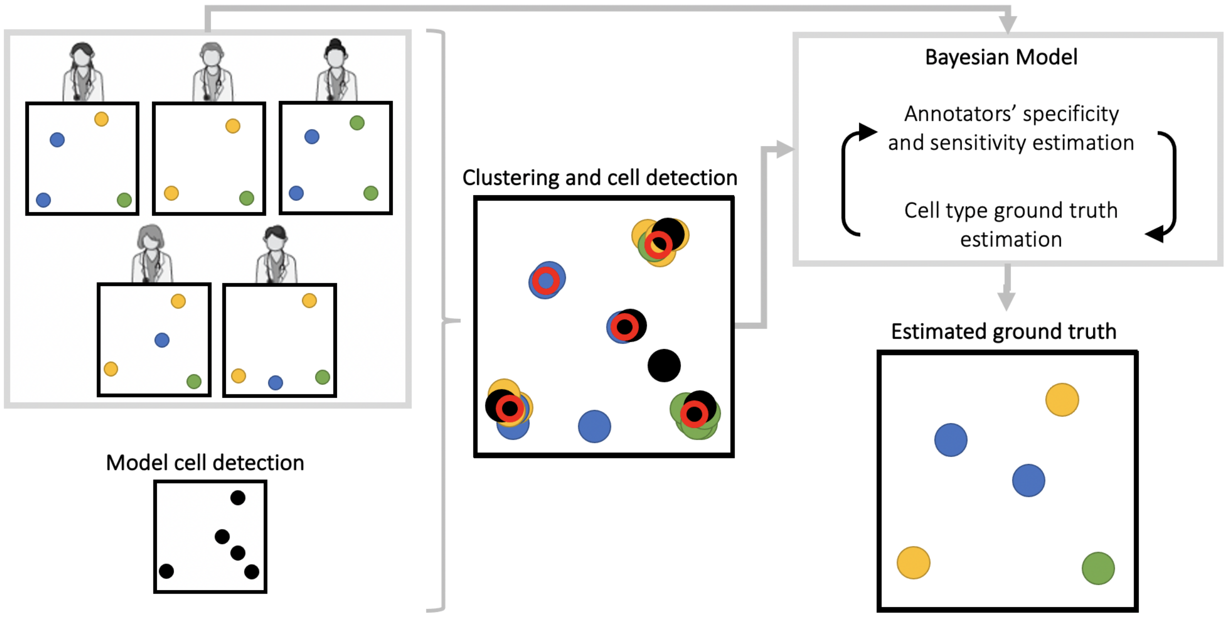

**Supplementary Figure 2. Frames analysis recall and precision.**

a

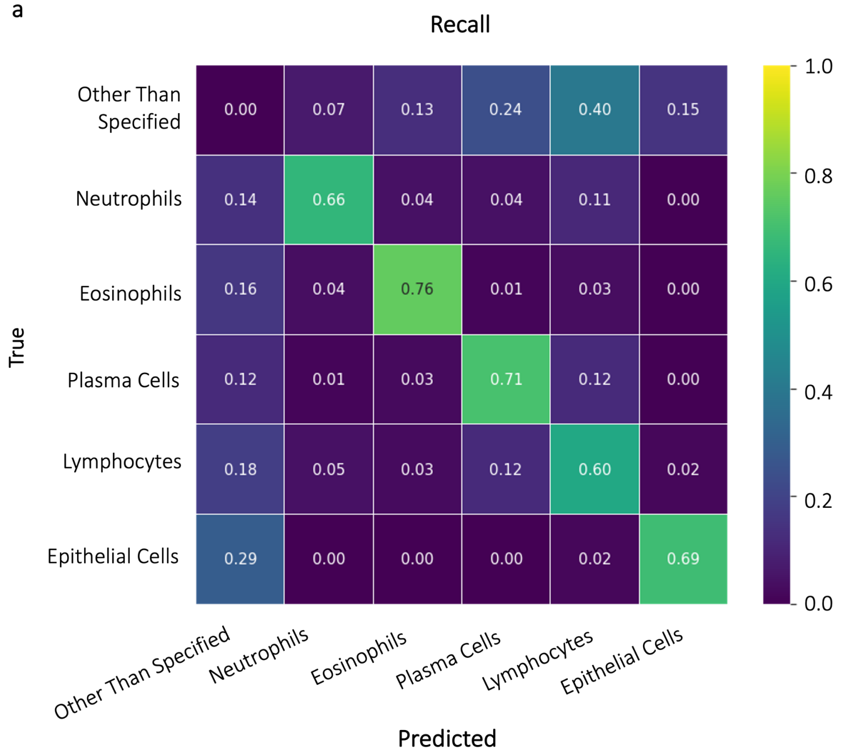


b

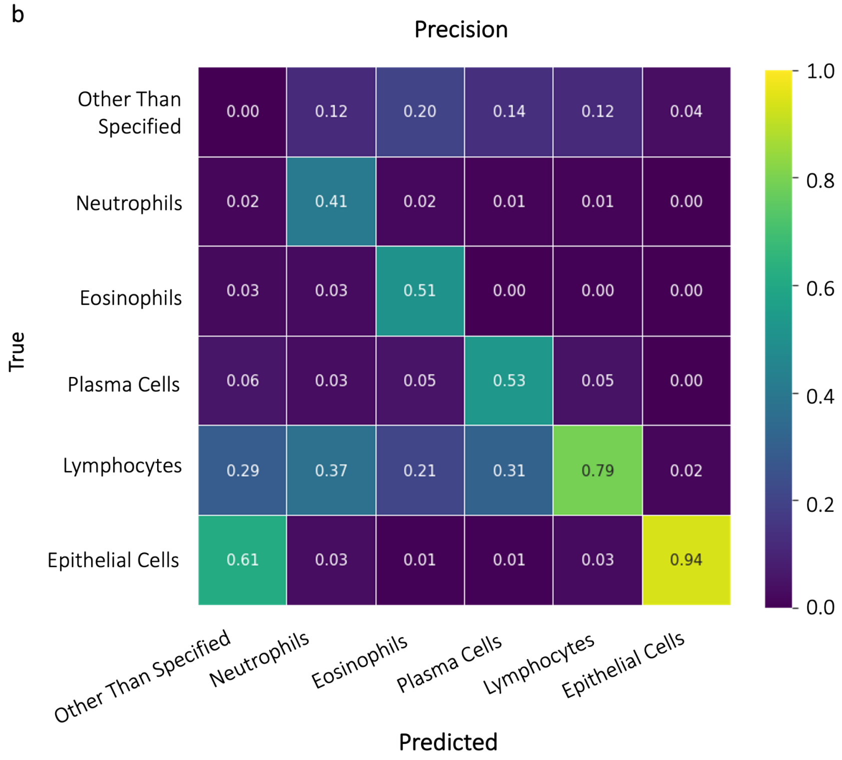

**Supplementary Figure 3. HIFs used in score determination by RCF.** The features are sorted by the permutation feature importance* performed on the held-out test set, which highlights the features with the most contribution to the model's generalization ability.

*Breiman, L. "Random Forests," *Machine Learning*, 45(1), 5-32, 2001.

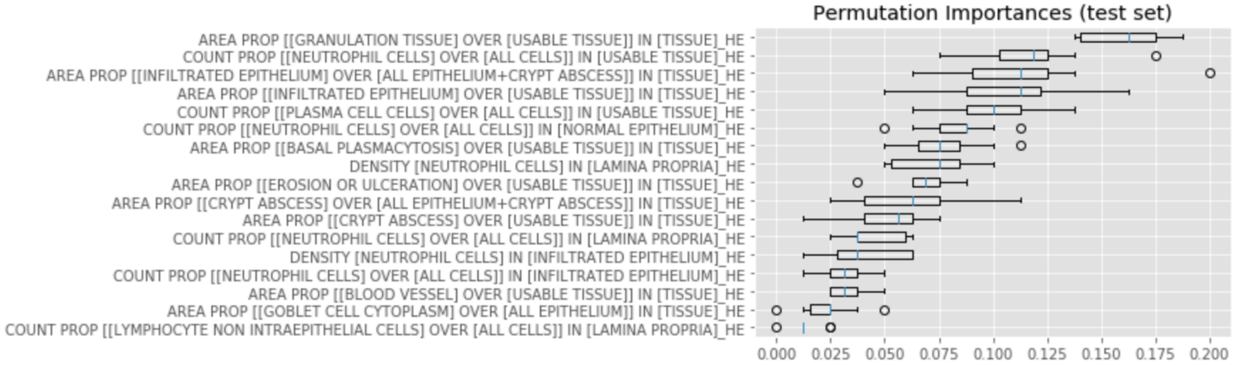
